# End Stage Kidney Disease in Cystic Fibrosis

**DOI:** 10.1101/2024.09.19.24313925

**Authors:** Mirjana Stevanovic, Todd A MacKenzie, Asha Zimmerman, Martha L Graber

**Affiliations:** Department of Microbiology and Immunology, Geisel School of Medicine at Dartmouth, Hanover, New Hampshire; Departments of Biostatistics and Medicine, Dartmouth Hitchcock Medical Center, and Geisel School of Medicine, Lebanon, NH; Department of Surgery, Dartmouth Hitchcock Medical Center, and Geisel School of Medicine, Lebanon, NH; Department of Medicine, Dartmouth Hitchcock Medical Center, and Geisel School of Medicine, Lebanon, NH

## Abstract

**Background:** Cystic fibrosis (CF) is a multisystem autosomal recessive disease caused by mutations in the cystic fibrosis transduction regulator (CFTR). The CFTR is expressed in all renal tubular segments. Tubular functional abnormalities as well as renal histologic changes are recognized in people with cystic fibrosis (CF). Chronic kidney disease (CKD) occurs in CF at greater frequency than in the non-CF population. The prevalence and characteristics of end stage kidney disease (ESKD) in the cystic fibrosis population have not previously been described.

**Methods:** We used data supplied by the US Renal Data System (USRDS) to compare persons with ESKD and CF with those with ESKD who did not have a diagnosis of CF. We used linear and logistic regression, propensity score-matched Kaplan-Meier survival curves, and log-rank tests.

**Results:** The prevalence of ESKD was approximately thirty times higher than predicted in the general US population. Diabetes was more frequently a diagnosis in people with CF, but was approximately half as frequently the primary cause of ESKD than in those without CF. Complications of transplantation, particularly of lung, were rarely the cause of ESKD in those without CF but were the second most frequent etiology of ESKD in people with CF. Drugs and medications, interstitial nephritis and acute kidney injury (AKI) were more frequent primary causes in those with CF. Hypertension, cystic kidney disease, and glomerulonephritis were less frequent in those with CF. Mortality and survival with ESKD were not significantly different between those with CF and ESKD and those with ESKD who did not have CF. Diabetes mellitus and complications of lung transplantation as the primary cause of ESKD, but not lung transplant status, were associated with significantly shorter survival with ESKD in people with CF.

**Conclusions:** People living with CF have a markedly higher prevalence of ESKD than the general United States population. The profile of primary causes of ESKD differed significantly from that of the non-CF population. Diabetes mellitus was approximately half as frequently the primary cause of ESKD in those with CF. Complications of solid organ transplant, particularly lung, were the second most frequent primary cause of ESKD in people with CF. Survival with ESKD was not significantly different between those who did and did not have CF. The diagnosis of diabetes mellitus, and complications of lung transplantation as the primary cause of ESKD, but not lung transplant status, were associated with significantly shorter survival with ESKD in people with CF. It is our aim that these findings will promote awareness of kidney disease in persons living with CF and will encourage prevention of acute kidney injury (AKI), early detection and intervention in CKD, and preemptive referral to nephrology and transplant centers.

## Introduction

CF is a multi-system disease caused by mutations in the cystic fibrosis transduction regulator (CFTR). CF affects between 30,000 and 40,000 people in the US^1,2^, and over 160,000 worldwide^3^. The CFTR is a cAMP-regulated transmembrane anion channel expressed in the epithelia of multiple organs including the skin, respiratory tract, reproductive tract, bowel, pancreas, and in all segments of the renal tubular epithelium^4,5^. In the kidney, the CFTR is implicated in the regulation of transport of sodium, chloride, bicarbonate, and potassium ions, regulation of urine osmolality, endosomal function^6,7^ renal development^8^, the development of kidney fibrosis^9^, and in cyst formation^10^. Although no specific renal phenotype other than nephrolithiasis is generally recognized in CF^11^, previous studies demonstrate increased susceptibility to chronic kidney disease (CKD) in persons with CF, and higher prevalence of CKD than in the general population^12^. The rate of CKD is reported to be 2.6% in adults with CF. Direct measures of glomerular filtration rate (GFR) demonstrate loss of kidney function, albuminuria and/or hyperfiltration in children and in up to 45% of adults with CF, with serum creatinine levels generally in the normal range for age. In addition to renal tubular dysfunction, disease-specific factors contributing to the relatively high rate of CKD are likely to include the very high lifetime prevalence of diabetes mellitus^13^, recurrent AKI during acute illness^14^, exposure to nephrotoxic drugs and medications in the management of infections and organ transplantation, and dehydration due to solute loss in sweat and the gastrointestinal (GI) tract. CKD of all etiologies progresses over months to years to ESKD requiring dialysis or kidney transplantation. Given the lack of population data concerning ESKD in CF, we sought to describe the prevalence, demographic characteristics, disease associations, and survival outcomes of people with CF and ESKD. We aim to enhance awareness of kidney disease in CF, and to encourage early detection and intervention in kidney disease in this patient population.

## Methods

To compare the predicted and actual number of individuals with both ESKD and CF, we used published data from the USRDS annual data report^15^, the Cystic Fibrosis Foundation Annual Data Report^1^, US census^16^, and a data request to the USRDS. To compare persons with ESKD with and without a diagnosis of CF, we used data linked to de-identified persons listed in the USRDS Patient and Hospitalization datasets during the five-year period January 1, 2014, to December 31, 2018. The ESRD Medical Evidence Report does not include CF as a diagnostic category. We relied upon comparisons between those with and without CF listed in the Hospitalization dataset.

The statistical analysis consisted of (i) estimation of the prevalence of CF in persons with ESKD with comparison to the general population, (ii) comparison of the characteristics at the time of incident ESKD between those with and without CF, (iii) comparison of the primary cause of ESKD between those and without CF, and (iv) estimations of the effect of CF on survival in individuals with ESKD. To compare the prevalence of CF in individuals with CF to the general population, we restricted to a younger age, < 44 years, and used prevalence data from the Cystic Fibrosis Foundation and published reports. Unless otherwise indicated all results are adjusted for age at ESKD incidence. To compare patient characteristics between those with and without CF, we used linear and logistic regression. We compared the primary cause of ESKD between CF and non-CF using logistic regression. To test the association between CF and outcomes of interest, we used multivariable logistic regression. Patient characteristics were selected for inclusion in these models based on forward and backward stepwise regression for each outcome. The stepwise model fit criterion was the Akaike Information Criterion. We compared survival between CF and non-CF, and between primary cause subgroups using Kaplan-Meier and log-rank tests while matching using propensity scores. In particular we used stratification/binning by propensity scores, matching for variables including incidence age, BMI, race and ethnicity, diabetes as the primary cause of ESKD, diabetes diagnosis, lung transplant complications as the primary cause of ESKD, and lung transplant status^17^.

Summary statistics for the racial and ethnic categories American Indian or Alaska Native, Asian, Native Hawaiian or Pacific Islander, and Other or Multiracial, were not included in these analyses because of small numbers of individuals within these categories with a diagnosis of cystic fibrosis. Some categories of primary cause of ESKD listed in the ESRD Medical Evidence Report were not included in these analyses.

## Results

### Prevalence of ESKD amongst those with and without CF

The 2016 USRDS annual data report listed 703,243 persons with ESKD, of whom approximately 6% (42,195) were under age 44 years^15^. The total US population in 2016 was 323,127,515, and the population under the age of 44 years in 2016 was 189,697,880^16^. We estimated the population prevalence of ESKD amongst US persons of all ages in 2016 to be 1:460, and of ESKD in US persons under the age of 44 in the same year was 1:4496.

To obtain an estimate of the frequency of ESKD in the CF population, we used estimates of the US population with CF in 2016. USRDS reported 236 US persons with diagnoses of both ESKD and CF in 2017 (USRDS data request). The CFF Patient Registry listed 29,577 persons with CF in 2016^1^. The CFF estimates capture of 85% of all persons living with CF in the US, yielding an estimated total number of persons with CF 34,796. Cromwell et al^2^ estimated the population prevalence of adults with CF in the years 1968 – 2020 to be 38,804 persons. The median age of those in the CFF registry in 2016 was 19 years; the median age at death of those in the CFF registry in 2016 was 29.6 years. Using these data, we estimated that the prevalence of ESKD amongst those with CF in 2016 was between 1:147 and 1:164.

### Comparison of persons with ESKD with and without CF

The USRDS Hospitalization data set listed 379 unique individuals with diagnoses of both ESKD and CF during the five-year period 1st January 2014 to 31st December 2018. In the same period there were 1,360,123 unique individuals with ESRD in the Patient dataset, and 954,203 individuals with ESKD who did not have a diagnosis of CF and appeared in both the Patient and Hospitalization datasets. To establish that those without CF listed in both the Patient and Hospitalization datasets were an appropriate control group, we performed univariable analyses of all variables used for comparison between CF and non-CF in this study to compare those without CF listed in the Patient dataset and those without CF listed in both the patient and hospitalization dataset. There were no significant differences detected between these groups in the characteristics we used for comparison between those with and without CF (data not shown).

### Characteristics of US individuals with cystic fibrosis and ESKD

People with CF and ESKD were on average 16.5 years younger than those without CF at ESKD incidence, 41.9 years vs 58.4 years (p < 0.001), had lower BMI by 6.93 (p < 0.001), and higher odds of being female, 51.2% vs 43.1% (OR 1.46; 1.19, 1.78; p < 0.001) (Table 1). Those with CF had higher odds of being identified as white, 87.3% vs 64.8% (OR 4.78; 3.53, 6.47; p < 0.001), and lower odds of being identified as black or African American, 10.8% vs 29.0% (OR 0.23; 0.17, 0.32; p < 0.001), or of Hispanic ethnicity, 7.9% vs 13.9% (OR 0.44; 0.31, 0.65; p < 0.001). People with CF and ESKD had higher odds than those without CF of having received care by a nephrologist prior to entry to the ESKD program, 59.6% vs 56.2% (OR 1.42; 1.07, 1.89; p = 0.01).

**Table 1:**
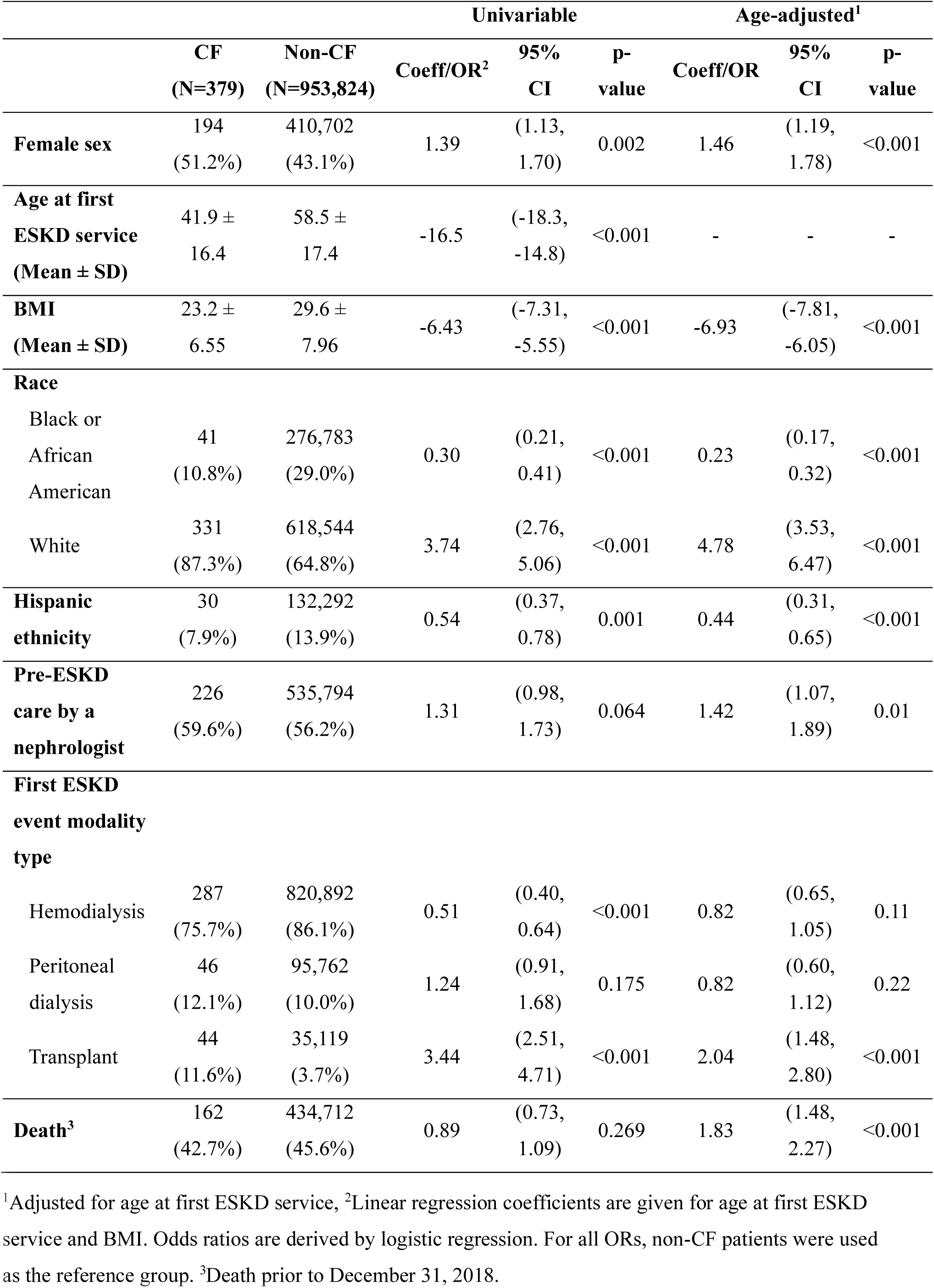
Demographic characteristics.

In age-adjusted univariable analyses, there was no significant difference between those with and without CF in the odds of having initiated ESKD care with peritoneal dialysis (OR 0.82, 0.60, 1.12; p = 0.22), or hemodialysis (OR 0.82, 0.65, 1.05; p = 0.22). In multivariable modeling controlling for incidence age, sex, BMI, race, ethnicity, and pre-ESKD care by a nephrologist, CF was associated with lower odds of having initiated ESKD with peritoneal dialysis rather than hemodialysis (OR 0.64; 0.44, 0.92; p = 0.02). In age-adjusted analysis, people with CF had higher odds of having initiated ESKD care with a kidney transplant vs dialysis (OR 2.04; 1.48, 2.80; p < 0.001). In multivariable modeling controlling for incidence age, sex, BMI, race, ethnicity, donor type, and pre-ESKD care by a nephrologist, those with CF were not significantly more likely to have initiated ESKD care with a kidney transplant (OR1.18; 0.77, 1.82; p = 0.44).

### Primary causes of ESKD

Diabetes was listed as a diagnosis in 63.6% of those with CF and 62.3% of those without CF (OR 1.44; 1.17, 1.78; p < 0.001) and was the primary cause of ESKD in 24.3% of those with CF and in 42.3% of those without CF (OR 0.56; 0.44, 0.71; p < 0.001) (Table 2). Of those with diabetes as the primary cause of ESKD, type 1 diabetes was identified in 34.8% of those with CF and in 11.0% of those without CF (OR 1.18; 0.82, 1.71; p = 0.37), and type 2 diabetes was identified in 65.2% of those with CF and 89.0% of those without CF (OR 0.45; 0.34, 0.59; p < 0.001). Of all those with diabetes as the primary cause of ESKD, those with CF had higher odds of being female than those without CF (OR 1.91; 1.26, 2.89; p = 0.01), were younger by 13.9 years (−16.7, −11.1; p < 0.001) and had lower BMI by 6.46 (−8.10, −4.82; p < 0.001) (data not shown). Hypertension was the cause of ESKD in 9.8% of those with CF and in 26.3% of those without CF (OR 0.42; 0.30, 0.58; p < 0.001). The ESRD Medical Evidence Report does not include CF as a diagnostic category, therefore no patients had CF listed as the primary cause of ESKD.

**Table 2:**
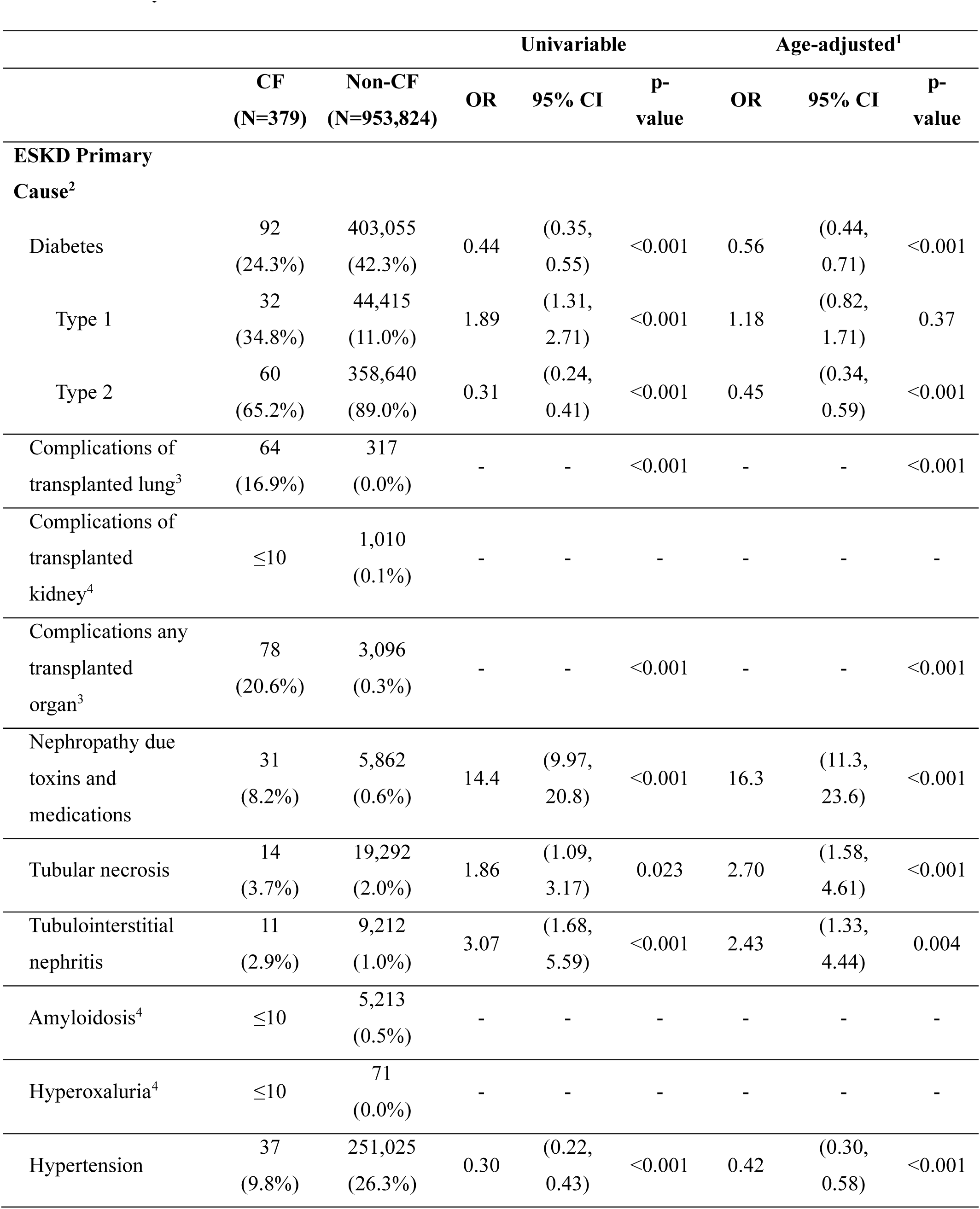
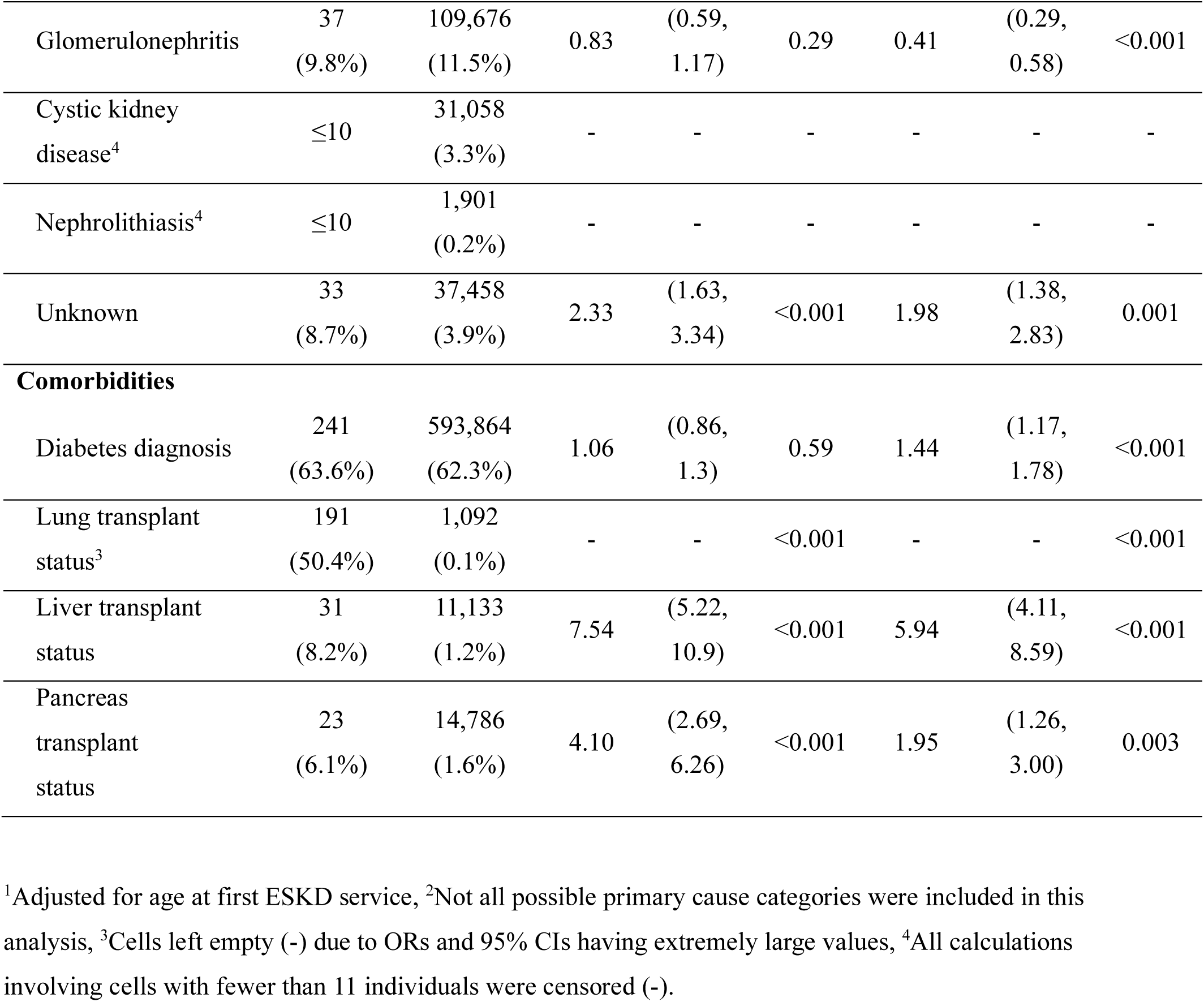
Primary causes of ESKD and comorbidities.

Complications of lung transplantation were the primary cause of ESKD in 16.9% of those with CF and 0% of those without CF (OR 682; 501, 929; p < 0.001). Complications of organ transplantation including lung were the primary cause of ESKD in 20.6% of those with CF and in 0.3% those without CF (p < 0.001). Lung transplant status was listed in 50.4% of those with CF and in 0.1% of those without CF (p < 0.001). Liver transplant status was listed in 8.2% of those with CF and in 1.2% of those without CF (OR 5.94; 4.11, 8.59; p < 0.001). Pancreas transplant status was listed in 6.1% of those with CF and in 1.6% of those without CF (OR 1.95; 1.26, 3.00; p = 0.01). Nephropathy due to drugs or other agents was the primary cause of ESKD in 8.2% of those with CF and in 0.6% those without CF (OR 16.3; 11.3, 23.6; p < 0.001).

### Mortality and survival

In age-adjusted univariable analysis, those with CF had higher odds of death occurring prior to the end of the study period, 42.7% vs 45.6% (OR 1.83; 1.48, 2.27; p < 0.001) (Table 1). In multivariable modeling controlling for age, sex, BMI, race, ethnicity, pre-ESKD care by a nephrologist, dialysis type, kidney transplant status, and donor type, those with CF did not have significantly greater odds of death prior to the end of the study period (OR 1.29; 0.88, 1.89; p = 0.19).

Median survival with ESKD was 12.6 years for those with CF and 11.8 years for those who did not have CF (p = 0.96) (Figure 1a). Those with CF who had diabetes as the primary cause of ESKD had median survival 5.58 years; those with CF who had a primary cause of ESKD other than diabetes had median survival of 13.4 years (p < 0.001) (Figure 1b). Comparing all with diabetes as the primary cause of ESKD, those with CF had median survival 5.58 years, and those who did not have CF had median survival 12.6 years (p < 0.001) (Figure 1c). For diabetes diagnosis (with or without diabetes as the primary cause of ESKD), those with CF had median survival 10.1 years, and those without CF had median survival 23.5 years (p< 0.001). Comparing all with complications of lung transplant as the primary cause of ESKD, those with CF had median survival of 12.6 years; those who did not have CF had median survival of 26.9 years (p < 0.001). For those with lung transplant status (with or without complications of lung transplant as the primary cause of ESKD) those with CF had median survival of 12.9 years and those who did not have CF had median survival of 11.7 years (p = 0.53) (Figure 1d)

**Figure 1:**
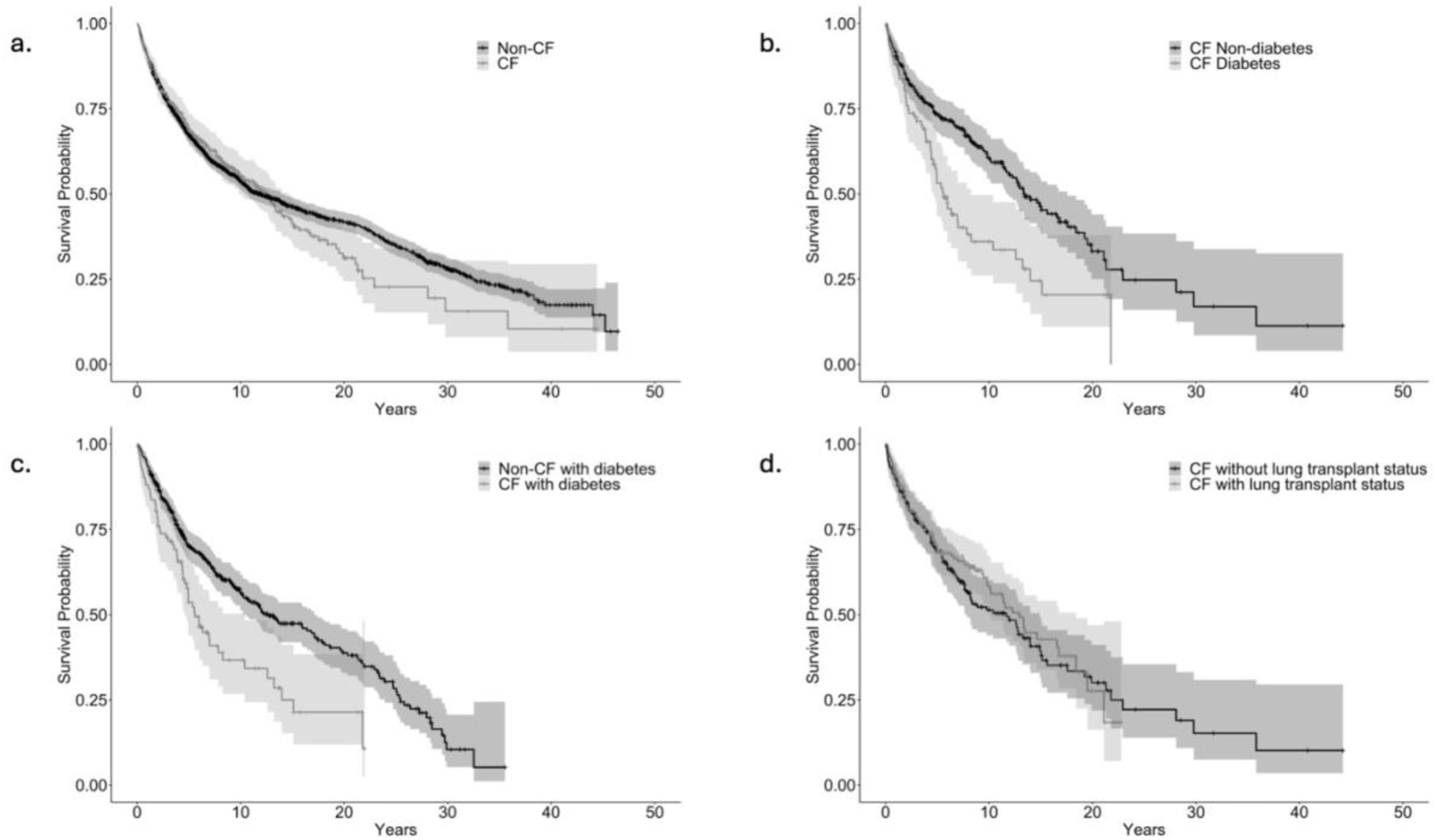
Survival of ESKD patients with and without CF, diabetes, and lung transplant. a. Overall survival of ESKD patients with CF, 12.6 years, and without CF, 11.8 years, (p = 0.96) with propensity score matching for age at first ESKD service, BMI, Black and White race, Hispanic ethnicity, diabetes diagnosis, diabetes as cause of ESKD, lung transplant status, and complications of transplanted lung as primary cause of ESKD. b. Overall survival of CF patients with diabetes as primary cause of ESKD, 5.58 years, or with a cause of ESKD other than diabetes, 13.43 years (p < 0.001), with propensity score matching for age at first ESKD service, BMI, Black and White race, and Hispanic ethnicity. c. Overall survival of patients with CF with diabetes as primary cause of ESKD with CF, 5.58 years, and patients without CF with diabetes as the primary cause of ESKD, 12.6 years (p < 0.001), with propensity score matching for age at first ESKD service, BMI, Black and White race, and Hispanic ethnicity. d. Overall survival of CF patients with lung transplant status, 12.9 years, and without lung transplant status, 11.7 years (p = 0.53) with propensity score matching for age at first ESKD service, BMI, Black and White race, Hispanic ethnicity, diabetes diagnosis, and diabetes as cause of ESKD.

## Discussion

As far as we are aware this is the first description of ESKD in people with CF. People with ESKD represented approximately 0.8% of all US persons living with CF^1^. In comparison, persons with ESKD under the age of 44 years comprised approximately 0.01% of the US population under the age of 44 years^16^. This may be an underestimate of the prevalence of ESKD amongst those with CF, because we were not able to determine from the available data how many people with CF and late stage or symptomatic CKD were not referred to a nephrologist, were not offered dialysis or transplantation if referred, or elected conservative care.

The average age of people with CF and ESKD in this study was 41.9 years, whereas the mean age of all US persons living with CF was 21.3 years, suggesting that this group may have both increased susceptibility to kidney disease and longevity allowing CKD to progress to ESKD over several years. Specific CFTR mutation classes could be relevant to the development and progression of ESKD. We were not able to classify CFTR mutation types in this study, and this is an area of particular interest for future investigation. The sex distribution in the general CF population is approximately equal, as was the sex distribution amongst those with CF and ESKD. This contrasts with the non-CF population with ESKD, who were more likely to be male. During the study period, an average of 4% of all people with CF in the US, and 11% of those with CF and ESKD, were identified as Black or African American. The racial disparity in the prevalence of ESKD in those with CF was similar to that recognized in the general ESKD population^18^, with the caveat that the prevalence of both CF and ESKD in those of African ancestry may be underestimated due to the observed underdiagnosis of CF in non-white ethnic groups^19^.

Diabetes mellitus was only half as frequently the primary cause in people with CF and ESKD, despite the very high lifetime rate of diagnosis of cystic fibrosis-related diabetes (CFRD) in people living with CF. CFRD is a non-autoimmune entity with features of both type 1 and type 2 diabetes and is a consequence of pancreatic inflammation and fibrosis and hormonal changes. The rate of CFRD increases with age, with current lifetime incidence of 50-80%^20^, compared with a lifetime risk of about one third in people without CF^21^. Diabetic nephropathy, retinopathy, and hypertension do occur in CFRD^22^, although the clinical significance of diabetic nephropathy in CFRD is not known. Both type 1 and type 2 diabetes mellitus can occur in people with CF. We could not distinguish individuals with CFRD from those with type 1 or type 2 diabetes in this study because CFRD is not specified by the Medicare eligibility form. We infer that at least a proportion of those with diabetes had CFRD because a higher proportion of those with CF than those without CF were classified as type 1. Taken together these results suggest that CFRD may carry a lower risk of nephropathy than endemic diabetes. Prospective studies would be required to determine the renal outcomes and risk of ESKD in CFRD. The diagnosis of diabetes, either as the primary cause of ESKD or as a comorbid diagnosis, was associated with significantly reduced survival with ESKD for those with CF compared both with those with CF without diabetes, and with those with diabetes who did not have CF. This is consistent with data that diabetes is associated with excess mortality in people with CF^23^, and is associated with higher all-cause mortality in the overall population with ESKD and diabetes^24^.

The second most frequently identified cause of ESKD in people with CF was complications of solid organ transplantation, particularly lung. Organ transplant complications were very infrequent causes of ESKD in people who did not have CF. Lung transplantation prolongs survival in those with advanced pulmonary complications of CF^25^. It is considered a last resort therapy, with only 2% of all US adults with CF in 2016 living with a lung transplant^1^. In contrast, of those with CF and ESKD half were living with a lung transplant. Renal complications are frequent following lung transplantation: AKI occurs in approximately half of all lung transplant patients, and nearly one in ten require dialysis^26^. The use of calcineurin inhibitors (CNI) in immunosuppression, together with increased tendency to dehydration and CF-specific tubular abnormalities^27^ all likely contribute to the higher risk of cumulative nephron loss. Kidney function declines more rapidly following lung transplantation in CF than those transplanted for other reasons^28^. Decreased GFR is documented in 35% of those with CF at two years following lung transplant, and in 58% at five years^29^. The relatively high rate of ESKD in this group may also be a consequence of serious lung exacerbations with AKI, and with factors independently associated with longevity with CF. Finally, selection bias may be present because those with CF who require lung, liver, or pancreas transplant are more likely to be followed in multidisciplinary transplant centers with experience in kidney transplantation.

Other primary causes of ESKD in those with CF reflect tubulo-interstitial injury. Tubular cell injury and subsequent interstitial fibrosis represent a common pathway in drug toxicity, interstitial nephritis, and hemodynamic AKI. Functional abnormalities described in CF include elevated urinary calcium and oxalate excretion, hypocitraturia^30,31^, persistent metabolic alkalosis^27^, and nephrolithiasis^11^, each of which may contribute to the risk of AKI and subsequent permanent nephron loss. Histological abnormalities described include tubulo-interstitial calcinosis in autopsies of infants and children, tubulo-interstitial injury and fibrosis, renal and systemic amyloidosis, and glomerular abnormalities^32^. Consistent with this, we found that AKI was present in the record of significantly more of those with CF than those without, and both tubular necrosis and tubulointerstitial nephritis were more frequent primary etiologies of ESKD in persons with CF. Nephropathy due to drugs and toxins was the etiology of ESKD in significantly more of those who had CF than those who did not. Specific agents could not be identified but are likely to have included CNIs and nephrotoxic antibiotics. Aminoglycosides are widely used in CF for both prophylaxis and treatment of pulmonary exacerbations. Despite the recognized nephrotoxicity of these agents, previous studies have been inconclusive concerning the contribution of cumulative aminoglycoside exposure to CKD in CF^33,34^. Less frequent etiologies of ESKD including amyloidosis and hyperoxaluria, albeit uncommon, were more frequent causes of ESKD in those with CF than in those without, consistent with the reported histologic and functional data. Perhaps surprisingly, given the relative frequency of kidney stones in those with CF, nephrolithiasis was a very infrequent cause of ESKD in this population. Both glomerulonephritis and cystic kidney disease were less frequently the primary cause of ESKD in those with CF than in those without CF.

These observations have important implications for awareness and management of kidney disease and kidney injury in people with CF. We describe a markedly increased prevalence of ESKD requiring dialysis or transplantation, pointing to the necessity of rapid and convenient measurements of kidney function, prevention of AKI, and early interventions to delay or prevent the progression of CKD to ESKD^34,35^. In turn, the observation that survival with ESKD for those with CF was comparable to that of those without CF, even while controlling for age and demographic and disease factors, prompts consideration of early referral to nephrology and transplant centers. A strategy of vigilance and active management of kidney disease will become more urgent as the CF population ages, and as new CF therapies - with uncertain impact on kidney function and CDK - become available^36,37^.

Unfortunately, detection of decreased GFR in CF is challenging for several reasons. AKI may be overlooked in the context of severe acute systemic illness. CKD may be missed in early stages because serum creatinine does not increase until significant nephron loss has occurred due to hypertrophy of remaining nephrons^38^, although this stage may be marked by albuminuria and the presence of urinary markers of tubular injury. Further, CKD is asymptomatic until advanced stages: the signs and symptoms of uremia including anemia, metabolic bone disease, fatigue, anorexia, and sarcopenia do not appear until 70-80% of nephrons have been lost and are frequently indistinguishable from those of chronic infection and nutritional deficiency related to pancreatic and gut dysfunction. The most critical obstacle to accurate detection and monitoring of kidney function in CF is that the standard formulae used for rapid estimation of GFR from serum creatinine and demographic or biometric data are inaccurate in adults or children with CF and may overestimate GFR^39^. Urinary markers of injury hold promise of specificity, but none are in current clinical use^40^. Given the current limitations, we recommend consideration of annual monitoring of renal function using 24-hour urine for creatinine, iothalamate clearance, or 99Tc DTPA renal scan, with urine microscopy, and quantitative analysis of urine total proteins. Identification of those at risk would allow initiation of strict blood pressure and glycemic control and the use of ACE inhibitors, angiotensin receptor blockers, SGLT2 inhibitors and GLP-1 agonists where appropriate, and timely referral to nephrology and transplant centers.

The impact of highly effective modulators of the CFTR, and of future therapies including gene editing on the incidence and progression of CKD to ESKD remains to be determined and is of great interest. This study provides baseline information about the prevalence and characteristics of ESKD in the current adult CF population. We have not attempted to address the potential impacts of newer therapies, which will manifest over years or decades. Highly effective modulators of the CFTR were approved for use in the US in 2019 and so would not be expected to have impacted the prevalence of ESKD during the period of data collection for this study. These agents may both preserve renal function and prolong life^37^ and are likely to impact both the prevalence of diabetes mellitus and the need for lung transplantation in those in whom they are both appropriate and available^41^.

### Limitations of this study

This is a retrospective study using de-identified data from large national datasets. We likely failed to account for all US persons with CF in this analysis: Cystic Fibrosis is not a specified diagnosis on the ESRD Medical Evidence Report (form 2728). As detailed in the methods section, we relied on comparisons between individuals in the USRDS Hospitalized data set with and without a diagnosis of CF. In addition, the Cystic Fibrosis Foundation patient registry includes most but not all US persons with CF.

## Data Availability

All data produced in the present study are available upon reasonable request to the authors

https://data.census.gov/table?q=US%20population%20in%202016&g=010XX00US&y=2016

## Disclaimer

The data reported here have been supplied by the United States Renal Data System (USRDS). The interpretation and reporting of these data are the responsibility of the author(s) and in no way should be seen as an official policy or interpretation of the U.S. government.

## Notes

### Competing Interest Statement

The authors have declared no competing interest.

### Funding Statement

This study was funded by a pilot study subaward to Dr Graber. The Prime grant is at Dartmouth College, PI Dean Madden DartCF P30DK117469

### Author Declarations

IRB review (copy on file with USRDS CC): The study was reviewed by the Dartmouth Hitchcock (Dartmouth Hitchcock Medical Center, Lebanon, NH 03756) IRB on 10.5.21. IRB ID STUDY02000769. The IRB determined that the study is not research involving human subjects as defined by DHHS and FDA regulations, and that IRB review is not required. IRB contact telephone 603 650-1846.

